# Effect of high-dose glucocorticoids on persistent opioid use 3 to 12 months after primary total hip or knee arthroplasty. Protocol for a target trial emulation using observational data from Danish registries

**DOI:** 10.1101/2023.10.31.23297517

**Authors:** Jens Laigaard, Robin Christensen, Claus Varnum, Martin Lindberg-Larsen, Troels Haxholdt Lunn, Ole Mathiesen, Søren Overgaard

**Affiliations:** Dept. of Orthopaedic Surgery and Traumatology, Bispebjerg Hospital, Copenhagen, Denmark; Dept. of Clinical Medicine, Faculty of Health and Medical Sciences, University of Copenhagen, Copenhagen, Denmark; Section for Biostatistics and Evidence-Based Research, the Parker Institute, Bispebjerg and Frederiksberg Hospital, Copenhagen, Denmark; Department of Clinical Research, University of Southern Denmark, Denmark; Department of Orthopaedic Surgery, Lillebaelt Hospital, Vejle, Denmark; Department of Regional Health Research, University of Southern Denmark, Denmark; Department of Orthopaedic Surgery and Traumatology, Odense University Hospital, Odense, Denmark; Department of Anaesthesiology and Intensive Care, Bispebjerg Hospital, Copenhagen, Denmark; Centre for Anaesthesiological Research, Department of Anaesthesiology, Zealand University Hospital Koge, Koege, Denmark

**Author notes:** Corresponding author’s contact information, Address: 2. floor, Nielsine Nielsens vej 6, 2400 Copenhagen NV, Denmark. **Funding:** No authors received external funding in relation to this study.

**Keywords:** Persistent postoperative pain, Pain transition, Hip replacement, Knee replacement

## Abstract

**Background:** Persistent postsurgical pain and opioid use after primary total hip and knee arthroplasty (THA and TKA) have major consequences for the patient and for society. High-dose perioperative treatment with glucocorticoids reduces inflammation and acute pain, both of which are associated with persistent postsurgical pain. We therefore hypothesise that routine treatment with glucocorticoids reduces the number of patients with persistent opioid use.

**Objective:** To determine if perioperative glucocorticoids for primary THA or TKA surgery, relative to no glucocorticoids, decreases the number of patients taking opioids in the period from 3 to 12 months after surgery.

**Design:** Target trial emulation trial with data from Danish national registries.

**Setting:** All departments of orthopaedic surgery in Denmark, from 1 January 2010 to 31 December 2020.

**Participants:** Patients with primary osteoarthritis undergoing primary THA or TKA, excluding presurgical users of glucocorticoids or insulin because these patients do not always receive the intervention.

**Intervention:** A single high-dose glucocorticoids (≥125 mg methylprednisolone or ≥24 mg dexamethasone) after induction of anaesthesia.

**Comparator:** No glucocorticoids during surgery.

**Allocation:** Patients operated at departments where treatment with high-dose glucocorticoids was standard of care at the time of surgery constitute the treatment arm, while patients operated at departments where high-dose glucocorticoids was not used serve as controls. Thus, all patients will be analysed according to their ‘allocation’, regardless of whether they received the treatment or not.

**Main outcome measures:** The primary outcome is number of persistent opioid users, defined as patients who redeem a prescription within at least two of the last three quarters during the first postsurgical year. The primary safety outcome is number of days alive and out of hospital within 90 days after surgery.

**Expectations:** These results will provide important evidence for or against the use of perioperative glucocorticoids in total hip and knee arthroplasty.

## Background

More and more patients are operated with total hip and knee arthroplasties (THA and TKA) due to primary osteoarthritis.^1–3^ The main indications are pain, reduced physical function and low quality of life.^4^ Internationally, about 9-20% of these patients continue to have moderate-severe persistent pain after surgery and approximately the same proportion remain opioid-dependent.^5,6^ Compared to the background population, patients with chronic pain have lower quality of life, more healthcare use, and higher risk of death.^7^ Opioid use amplifies these risks even further.^8^

Pain and inflammation in the days following surgery is associated with higher risk of persistent postsurgical pain.^9,10^ This association may be explained by upstream changes in the central nervous system (i.e. central sensitisation) caused by intense nociceptive stimuli from tissue damage.^10,11^ Glucocorticoids have marked anti-inflammatory and thereby also analgesic effects,^12^ which could impede these nociceptive stimuli and thereby reduce the risk of persistent postsurgical pain and opioid use.

After two Danish trials showed short-term benefits,^13,14^ high-dose glucocorticoids were implemented as routine treatment for THA and TKA at all Danish orthopaedic departments during the 2010s.^15^ However, due to concerns at single centres about adverse effects, e.g. increase risk of infection, this implementation happened gradually.^15^

## Rationale

Now, administration of glucocorticoids during surgery is an established treatment following THA and TKA in Denmark. We consider it important to investigate if the routine use of high-dose glucocorticoids reduces the risk of persistent opioid use after THA and TKA. We will therefore emulate a stepped wedge cluster-randomised trial with observational data, to investigate the long-term effects of routine use of high-dose of glucocorticoids for patients undergoing primary THA or TKA (Table 1). We hypothesise that glucocorticoids are superior compared to no treatment.

**Table 1:**
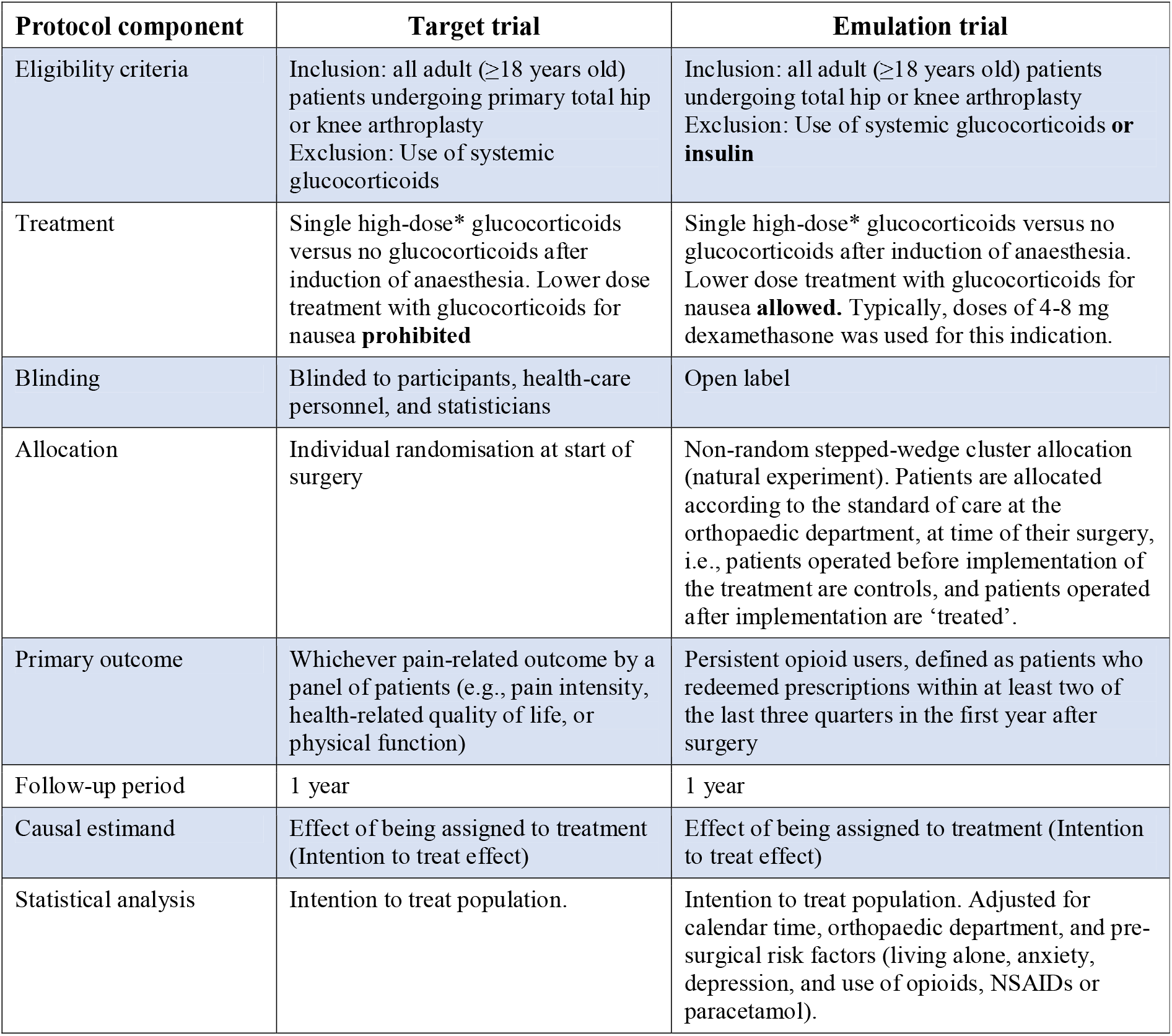
Differences between the target trial and the emulated trial. NSAID = non-steroid anti-inflammatory drug. *High-dose is defined as ≥ 24 mg dexamethasone or equipotent doses of other glucocorticoids.

## Objectives

Our primary effectiveness objective is to compare the effect of routine use of high-dose glucocorticoids (≥125 mg methylprednisolone or ≥24 mg dexamethasone) for primary THA or TKA surgery, relative to no or low dose of glucocorticoids, on the number of patients taking opioids in the period from 3 to 12 months after surgery.

As secondary effectiveness objectives, we will also assess the effect of routine use of high-dose glucocorticoids for primary THA or TKA surgery, relative to no or low dose glucocorticoids, on the number of patients taking prescription non-steroid anti-inflammatory drugs (NSAIDs) or paracetamol in the period from 3 to 12 months after surgery.

The safety objective is to compare the effect of routine use of high-dose glucocorticoids for primary THA or TKA surgery, relative to no or low dose glucocorticoids, on the number of days alive and out of hospital within 90 days after surgery.^16,17^

## Methods

This protocol is written according to the SPIRIT 2013 statement.^18^

### Study setting

All residents in Denmark have equal access to free-of-charge healthcare financed by general taxes. Each year, around 10,000 and 7,000 Danish patients will have a primary THA or primary TKA, respectively, due to primary osteoarthritis.^1,2^ Enhanced Recovery After Surgery (ERAS) guidelines are widely implemented in Denmark and half of the patients are discharged home the day after surgery.^1,2,19^

### Eligibility criteria

All eligibility criteria will be assessed at time of surgery. All adults (≥18 years old) who underwent elective, unilateral THA or TKA due to primary osteoarthritis between 1 January 2010 and 31 December 2020 are eligible. A patient can only enter the study once, meaning that patients undergoing more than one THA or TKA procedure, will only be analysed for the first entry. Patients with insulin dependent diabetes mellitus and patients receiving systemic glucocorticoids treatment will be excluded, as the intervention is sometimes precluded for these patients.^12^ Patients who underwent one-stage bilateral surgery are also excluded, as these procedures are rare.

### Intervention

The intervention is routine use of a single high-dose of glucocorticoids given after induction of anaesthesia. For each hospital, patients who underwent surgery before implementation of glucocorticoids serve as controls, while patients operated after implementation constitute the treatment arm, regardless if they received the treatment or not.^20,21^ The intervention was given along with other analgesic interventions that were routinely given to THA and TKA patients in Denmark during the study period. These include paracetamol, NSAIDs, opioids, and, for TKA patients, local infiltration analgesia.

### Comparator

The comparator is no administration of glucocorticoids during anaesthesia. Patients in the control arm may have received a lower dose of glucocorticoids for postsurgical nausea and vomiting. Typical anti-emetic doses are 4-8 mg dexamethasone.^22^

### Outcomes

The primary outcome is the number of persistent opioid users, defined as patients who redeemed prescriptions within at least two of the last three quarters during the first year following surgery.^23,24^ The first postsurgical quarter is excluded because the ICD-11 defines chronic postsurgical pain as pain “persisting beyond the healing process, i.e. at least 3 months after surgery”.^24^ We choose to use two redeemed prescriptions in separate quarters to ensure that the patient is in fact taking the medication.^25^ A list of the Anatomical Therapeutic Chemical (ATC) codes of the medication types used in the study can be found in Appendix 1 The number of persistent opioid users is summarised as n (% of all patients).

Secondary outcomes are:

1) Number of prescription-NSAID users, assessed in the same way as the primary outcome and summarised as n (% as all patients).
3) Number of prescription-paracetamol users, assessed in the same way as the primary outcome and summarised as n (% as all patients). The number of NSAID/paracetamol users constitute proxy measures for persistent postsurgical pain.
3) Days alive and out of hospital within 90 days of surgery.^16,17^ This is a composite outcome of death, length of hospital stay, and re-admissions. Higher numbers represent better outcomes and patients who died within the first 90 days after surgery will be assigned a value of zero. The outcome incorporates typically assessed safety outcomes, including readmissions due to re-operation, infections, thromboembolic events, and other serious adverse events. Days alive and out of hospital within 90 days is summarised as mean (SD). In our opinion, this is a pragmatic, patient-centred outcome that can be obtained reliably from the Danish registries.

### Assessment of variables

The exclusion criteria are assessed up to one year before surgery, i.e., patients who redeemed a prescription for systemic glucocorticoids or insulin in the year before surgery will be excluded from the study (Figure 1).

**Figure 1:**
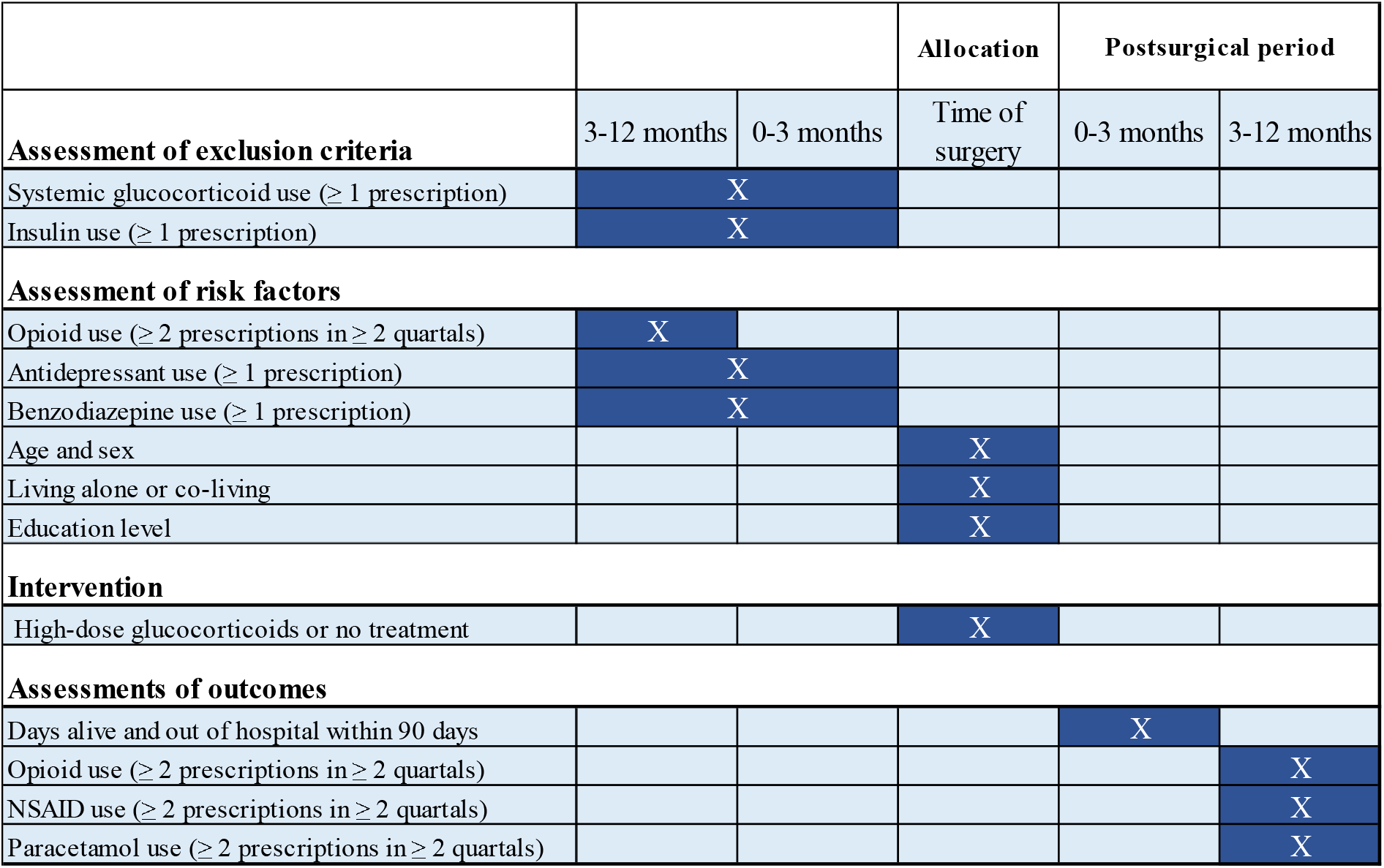
Participant timeline. NSAID = non-steroid anti-inflammatory drug.

All included patients are observed from one year before surgery to one year after surgery. In the year before surgery, important presurgical risk factors for persistent postsurgical pain are assessed. These include whether patients have redeemed a prescription of antidepressants or benzodiazepines in the year before surgery.^26^ Moreover, presurgical opioid use is assessed from 3 to 12 months before surgery and defined as redeemed prescriptions within at least two of the first three quarters during the year before surgery.^27^ We excluded the last quarter up to surgery from the definition of presurgical opioid use for two reasons. First, opioid prescriptions in this quarter may not reflect current use, but rather preparation for the upcoming surgery. Second, a study found that opioid use limited to this quarter was not associated with worse outcomes.^27^

Other risk factors will be assessed at time of surgery, including age, sex and living alone or co-living. As described in the ‘outcomes’ section, all outcomes are assessed in the first year following surgery.

### Sample size and power considerations

Because of similarities in data structure, we calculated the power of the study as were it a stepped-wedge cluster-randomised trial.

From 2010 to 2020, approximately 127,000 and 99,000 Danish patients underwent THA and TKA surgery, respectively.^1,2^ We do not expect to include all patients operated during the study period, mainly due to limitations in access to health care records. Thus, the sample size calculation is based on 120,000 analysed patients distributed evenly between the two arms and with 15 clusters/steps. (Figure 2). All clusters and steps were considered of equal size to simplify the calculation. The calculation is based on an intra cluster coefficient (ICC) of 0.05 and a cluster auto correlation (CAC) of 1.^28^ With estimated incidence of 10% and 11% persistent opioid users in the treatment and control group, respectively, the study has 90% power to obtain statistical significance at significance level of 0.05. We considered a 10% relative difference the minimal important difference.^29^ This calculation was performed in R with the swCRTdesign package (version 3.3).^30^

**Figure 2:**
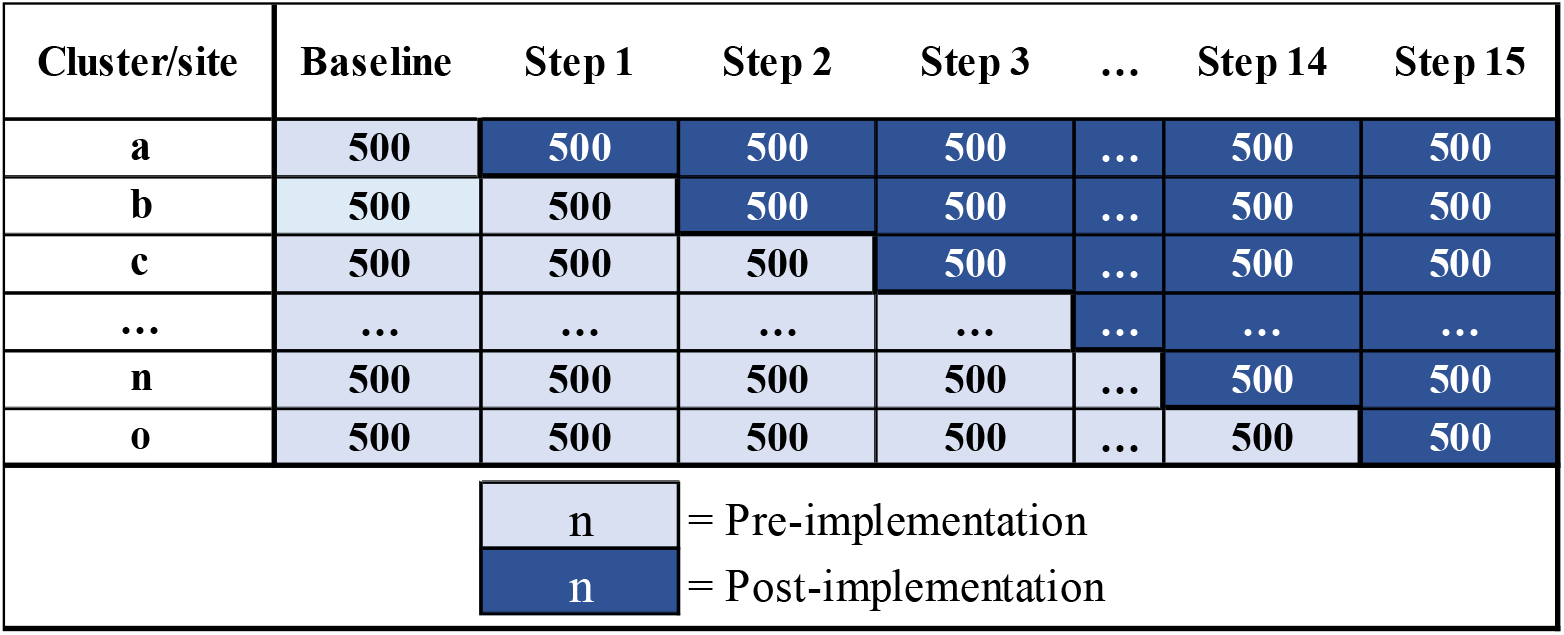
Schematic representation of the trial design with fictional numbers used for the sample size calculation. Each step represents the time point where a department starts treating patients with high-dose glucocorticoids. Clusters/sites and steps will have different sizes and lengths in the actual dataset.

### Recruitment

Patients will be identified through the Danish Hip and Knee Arthroplasty Registries. These high-quality registries have had coverages well over 90% of THA and TKA operations through the study years and comprise data reported by the operating orthopaedic surgeons.^31–33^ These patients are screened for eligibility with data from the Danish National Prescription Registry on systemic glucocorticoid or insulin use.^34^ Patients’ flow through the study will be presented in a attrition diagram (Figure 3).

**Figure 3:**
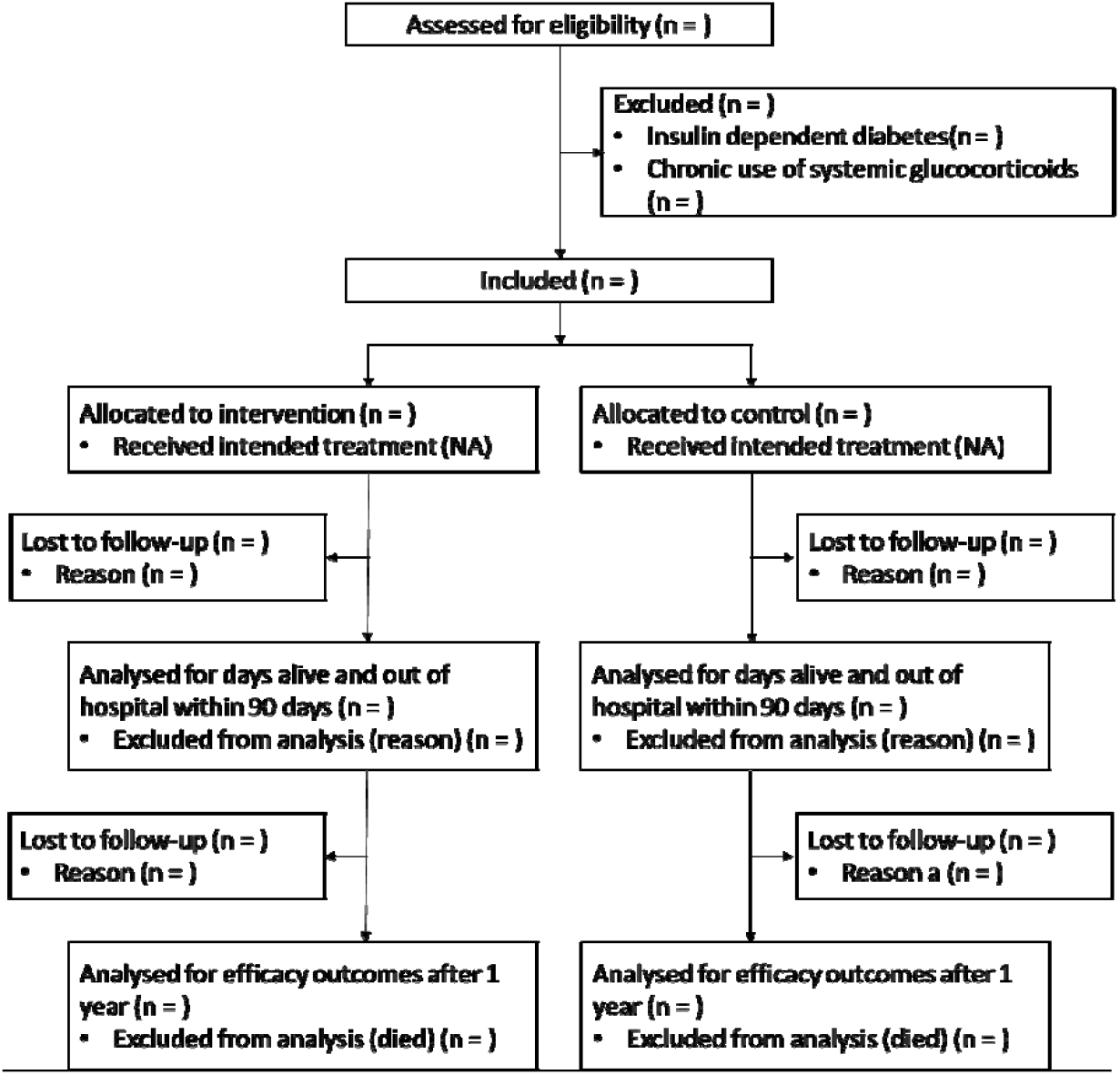
Attrition diagram to be presented in the paper. NA = Not available.

### Assignment of interventions

Treatment allocation is non-random and open label. Patients operated at departments where high-dose glucocorticoids were part of the standard analgesic regimen at the time of surgery constitute the treatment arm. Patients operated at departments where high-dose glucocorticoids were not routine at the time of surgery are assigned the control arm. Thus, to classify each patient into the control or the treatment arm, we must identify if the hospital routinely administered high-dose glucocorticoids at the time of surgery. For each hospital, if the exact date that the local guideline changed is unavailable, we will hand-search patient files to evaluate when this change in treatment occurred.^15^

Historically, the orthopaedic departments in Denmark have adopted different treatments at different times because surgeons have interpreted the evidence differently.^15,35^ In our experience, concerns about postsurgical infections and effect of treatment were the main cause of delayed implementation of glucocorticoids at many departments. It is possible that high volume departments or university hospitals systematically implemented the treatment before smaller departments. Therefore, we will report the observed “allocation” sequence in the resulting paper, with the total number of procedures performed in each department (Table 2).

**Table 2:** Fictional, chronological list of implementation of routine administration of high-dose glucocorticoids for total hip and knee arthroplasty in Denmark. THA = total hip arthroplasty. TKA = total knee arthroplasty.

| Site/department | Implementation of high dose glucocorticoid treatment | Average volume 2010-2020 (surgeries per year) |  |
| --- | --- | --- | --- |
|  |  | THA | TKA |
| Department a | October 1, 2013 | 311 | 199 |
| Department b | October 7, 2013 | 421 | 313 |
| Department c | October 8, 2013 | 65 | 45 |
| ... | ... | ... | ... |
| Department n | June 6, 2017 | 201 | 133 |
| Department o | June 6, 2017 | 145 | 97 |

### Data collection

Baseline data, including age, sex, type of anaesthesia, surgical access, use of local infiltration analgesia etc., will be obtained from the Danish Hip and Knee registries.^32,33^

The Danish Health Data Authority (www.sundhedsdatastyrelsen.dk/da/english) will supply data on dispensed prescriptions from the Danish National Prescription Registry, which contains all dispensed prescription medications in Denmark.^34^ Paracetamol and low-dose NSAIDS are available over-the-counter in Denmark, but since 30 September 2013, packs of more than 20 tablets have required a prescription. Opioids has only been available with a prescription throughout the study period.

Patients’ education level and whether patients are living alone or co-living will be assessed with data from Statistics Denmark (www.dst.dk/en). Please find the detailed list of data sources in Appendix 2. All residents in Denmark have a unique 10-digit personal identification number which allows us to link data from different registries.

### Data management

All eligible patients are identified from the Danish Hip and Knee Arthroplasty Registry and downloaded in an encrypted fashion to a drive with logged entry.^32,33^ These data are uploaded to Statistics Denmark’s secure online research platform in order to link with data from the Danish National Prescription Registry and .^34^ All data handling will be performed in this remote-access environment, which can only be accessed with the corresponding author’s digital ID. Within this environment, R statistical software with packages tidyverse, epitools, lme4, and marginaleffects and will be used for data handling and analysis.^36–40^

### Statistical methods

We will analyse the effect a single high-dose of glucocorticoids in two ways; 1) a crude, unadjusted analysis, 2) a pre-defined model adjusted for important pre-exposure variables using stabilised inverse probability of treatment weighting (SIPTW).^10,41^

1. The crude analyses of binary outcomes (i.e., persistent opioid/NSAID/paracetamol use) are conducted with the Fischer’s exact test and reported as relative and absolute risk differences with 95% confidence intervals (calculated assuming binomial distribution). The crude analyses of continuous outcomes (i.e., days alive and out of hospital within 90 days) are conducted with a welch t-test and reported as mean differences with 95% confidence intervals.
2. The primary analyses will be adjusted for department, calendar time, and known risk factors (Figure 4). First, we will apply a logistic mixed-effects model, using the lme4 package^39^, to generate propensity scores^42^ from the following variables:
  - Department (categorical variable), included as a random effect
  - Calendar time (continuous variable), included as a fixed effect
  - Age (continuous variable), included as a fixed effect
  - Sex (binary variable), included as a fixed effect
  - Education level (categorical variable), included as a fixed effect
  - Living alone or co-living (binary variable), included as a fixed effect
  - Type of surgery (binary variable: THA/TKA), included as a fixed effect
  - Presurgical use of antidepressants (binary variable: yes/no), included as a fixed effect
  - Presurgical use of benzodiazepines (binary variable: yes/no), included as a fixed effect
  - Presurgical use of opioids (binary variable: yes/no), included as a fixed effect

**Figure 4:**
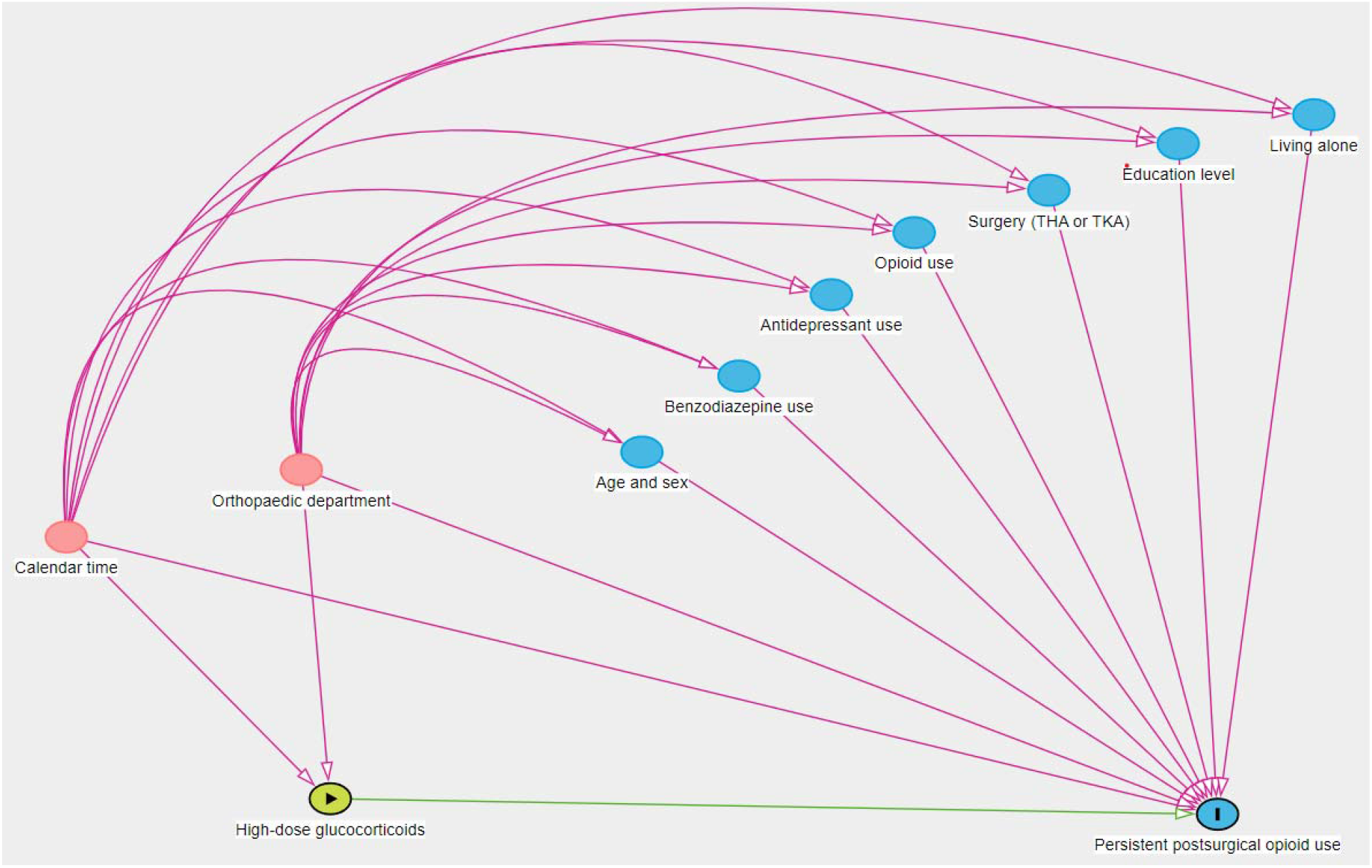
Directed Acyclic Graph (DAG).^43^ Intervention in green. Ancestors of outcome in blue. Ancestors of intervention and outcome (confounders) in red. THA = total hip arthroplasty. TKA = total knee arthroplasty.

Each unit is given weight by the inverse probability of being in their respective arm (treatment/control). That is, patients in the treatment arm are assigned weights as (propensity score)^-1^, while controls are assigned (1 - propensity score)^-1^. Hence, patients in the treatment arm with low propensity scores and controls with high propensity scores are given more weight. Finally, these weights are ‘stabilised’ by multiplying each unit’s weight by the proportion of units in their treatment group.^41^

These weights are used to calculate the average marginal estimates with the marginaleffects package in R.^40^ Again, binary outcomes are reported as mean relative and absolute risk differences with 95% confidence intervals, while continuous outcomes are reported as mean differences with 95% confidence intervals. The minimal important difference was considered a 10% relative reduction in opioid users from 3 to 12 months after surgery.^29^ P-values below 0.05 are considered statistically significant.

### Loss to follow-up and Missing data

Generally, no data will be imputed. Patients who died in the first year following surgery are analysed for days alive and out of hospital within 90 days, but not for efficacy outcomes (number of opioid/NSAID/paracetamol users). As 1-year mortality following THA and TKA is low, we do not believe this will influence the results. Some patients may have purchased opioids illegally and certainly some patients bought NSAIDs or paracetamol over the counter. Unless these patients also redeemed prescriptions for these medications, they will be analysed as non-users.

### Monitoring

Since this is an observational study, there is no need for a data monitoring committee or audits.

### Ethical considerations

Since the study relies solely on data from national registries, consent from patients and approval from the ethics committee are not needed. Following national data protection regulations, the project appears on the regional research listing (Privacy) with identifier P-2023-16. We obtained permission to access patient files from the Centre for Health, Capital Region of Denmark (identifier R-23002069). Data will be handled confidentially and only the study authors will have access.

### Reporting of results

The resulting paper will be reported according to the CONSORT 2010 statement with the extension for stepped wedge cluster randomised trials.^20,21^ Generally, we will report the results as would we report a stepped-wedge cluster randomised trial, including a diagram of participants flow through the trial. We will seek to make the paper freely available as preprint at www.medrxiv.org or through open-access publication. Moreover, the results will be presented at meetings and conferences, predominantly for surgeons, anaesthetists, pain specialists, and other health-care personnel working with this patient group.

## Discussion

This registry-based target trial will assess the long-term effects of a single high-dose glucocorticoids for THA and TKA surgery. Although observational, we believe that the current design is well-suited to confidently answer the research question.

Though a randomised trial would be preferable, our study has several advantages. First, it is possible to obtain data on a large, unselected cohort relatively fast. While we do impose a few exclusion criteria, our cohort is more likely to reflect the actual surgical population than participants in a randomised trial.^44^ Second, we use routinely collected prescription data that are not biased by knowledge of allocation (i.e., no Hawthorne effect).^45^ Third, we did not use the actual treatment status to categorize the patients into treatment groups. Instead, patients are allocated based on the *intended* treatment, i.e., the orthopaedic departments’ guideline. Therefore, the treatment allocation is not confounded by indication.^44,46,47^

Nevertheless, observational studies do carry biases that are accounted for in intention-to-treat analyses of randomised trials. In our study, there is a risk that the observed effect, or lack of effect, is simply due to temporal changes in treatment and prescription patterns, although we do adjust for calendar time in the statistical analyses. Still, it is not possible to control for co-implementation of other interventions (e.g., prescription of smaller opioid pack sizes).

We chose to adjust for known presurgical risk factors, or proxy-measures of risk factors, that we could reliably obtain from the Danish registries.^48^ To avoid problems with reverse causality, we chose not to adjust for risk factors that are determined after the intervention was given, such as anaesthesia, surgical complications, surgery length, acute postsurgical pain, and inflammation.^44^ Still, some important presurgical risk factors were not included, at least not directly. For example, smoking status and genetic disposition are known risk factors, but we are not able to include them in the analyses because neither are available from the registries in Denmark.^10^ Also, we did not include employment status in the propensity score model, as most patients operated with THA or TKA for primary osteoarthritis are retired at time of surgery. Lastly, psychological factors including anxiety, depression, and pain catastrophising are not included directly in the analyses, in part because they are difficult to assess from registry data. Instead, we included presurgical use of antidepressants and benzodiazepines, which are prescribed for various psychiatric disorders as well as patients with chronic pain.^49^

Overall, we believe this study will provide reliable and robust answers to the study objectives.

## Data Availability

All data used for this study is publicly available from
1) The Danish Clinical Quality Program for the National Clinical Registries at www.RKKP.dk
2) The Danish Health Data Authority at www.sundhedsdatastyrelsen.dk/da/english
3) Denmark Statistics at www.dst.dk/en

## Contributions

JL, TL, OM and SO conceived the idea for the protocol. JL drafted the first version of the protocol. All authors revised the manuscript and take responsibility for the content.

### Appendix 1 - Anatomical Therapeutic Chemical (ATC) codes of medications used in the study

**Antidepressants**: ATC codes N06A, excluding N06AX12 (bupropion)

**Benzodiazepines**: ATC codes N05BA

**Glucocorticoids**: ATC codes H02

**Insulin**: ATC codes A10A

**Non-steroid anti-inflammatory drugs (NSAIDs)**: ATC codes M01A, excluding M01AX (Other antiinflammatory and antirheumatic agents, non-steroids).

**Opioids**: ATC codes N02A, ATC code: R05DA04 (codeine) and ATC code N07BC02 (metadone).

**Paracetamol:** ATC code N02BE01

### Appendix 2 – Detailed data sources

#### 1) Danish Hip Arthroplasty registry (www.danskhoftealloplastikregister.dk/en/dhr/)

For patients who underwent Total Hip Arthroplasty, the following variables are assessed:

- Age
- Sex
- Date of surgery
- Department.
- In addition, baseline surgical data (surgical access, use of cement, etc.) are assessed.

#### 2) Danish Knee Arthroplasty registry (www.rkkp.dk/kvalitetsdatabaser/databaser/dansk-knaealloplastik-register)

For patients who underwent Total Knee Arthroplasty, the following variables are assessed:

- Age
- Sex
- Date of surgery
- Department.
- In addition, baseline surgical data (surgical access, use of cement, use of local infiltration analgesia etc.) are assessed.

#### 3) Danish Health Data Authority (www.sundhedsdatastyrelsen.dk/da/english)

For all patients, prescription redemption data (including CPR number date, number of packages, Anatomical Therapeutic Chemical (ATC) code, number of defined daily doses per package) for the following medication types and periods are assessed:

- Antidepressants – from 365 days before surgery and up to the date of surgery.
- Benzodiazepines – from 365 days before surgery and up to the date of surgery.
- Non-steroid anti-inflammatory drugs (NSAIDs) – from the date of surgery to 365 days after surgery.
- Opioids – from 365 to 90 days before surgery AND from 90 to 365 days after surgery
- Paracetamol – from the date of surgery to 365 days after surgery.

#### 4) Statistics Denmark (www.dst.dk/en)

For all patients, the following variables are assessed:

- Highest education attained, assessed by categorising the variable (ALMFSP) into categories according to the International Standard Classification of Education (ISCED) (https://www.dst.dk/da/TilSalg/Forskningsservice/Dokumentation/hoejkvalitetsvariable/hoejst-fuldfoerte-uddannelse/almfsp)
- Living alone or co-living, assessed by categorising the ‘family type’ (FAMILIE_TYPE) variable (https://www.dst.dk/da/TilSalg/Forskningsservice/Dokumentation/hoejkvalitetsvariable/familier/familie-type).

Both variables are classified as high-quality variables by Statistics Denmark, and undergo quality assurance internally and by external auditors.

